# Deep Learning-Based Automated Echocardiographic Measurements in Pediatric and Congenital Heart Disease

**DOI:** 10.64898/2026.02.06.26345782

**Authors:** Platon Lukyanenko, Sunil Ghelani, Yuting Yang, Bohan Jiang, Timothy Miller, Peter Higgins, Manouk Kirakosian, Kaitlyn Tracy, Janet Kane, David Harrild, John Triedman, Andrew J. Powell, Tal Geva, William G. La Cava, Joshua Mayourian

## Abstract

**Background:** Echocardiography (echo) is a cornerstone of pediatric cardiology, yet access to expert interpreters is limited worldwide, particularly in low-resource and rural settings. Artificial intelligence (AI) offers a mechanism to broadly deliver expert-level precision and standardize measurements, yet AI for comprehensive automated measurements in pediatric and congenital heart disease (CHD) echo remains underdeveloped.

**Methods:** We created EchoFocus-Measure, an AI platform that automatically extracts 18 quantitative parameters and 10 qualitative assessments from full echo studies. The method extends a multi-task, view-agnostic architecture (PanEcho) with a study-level transformer to prioritize diagnostically informative views. Training (80%) and internal testing (20%) were performed on echos from Boston Children’s Hospital (BCH), with external evaluation on outside referral studies. Left ventricular ejection fraction (LVEF) was the primary endpoint.

**Results:** The internal cohort included 11.4 million videos from 217,435 echos (60,269 patients; median age 8.5 years; median LVEF 61%), and external validation encompassed 289,613 videos from 3,096 echos (2,506 patients; median age 3.5 years; median LVEF 62%). For LVEF, EchoFocus-Measure exhibited a median absolute error (MAE) of 2.8% internally and 3.8% externally, maintaining accuracy across infants (MAE 3.2%) and complex CHD lesions (e.g., MAE 4.0% for L-loop transposition of the great arteries). EchoFocus-Measure improved upon the PanEcho benchmark (MAE 7.5% for infants; 13.1% for L-loop transposition). Discrepant case (>50^th^ percentile error) adjudication of LVEF demonstrated that model errors (MAE 2.4%) were within human variability (MAE 3.7%). For qualitative measures, EchoFocus-Measure performed well internally (AUROC 0.88-0.95) and modestly externally (AUROC 0.73-0.86). Explainability analyses highlighted model focus on clinically appropriate echo views for LVEF estimation (apical four-chamber, parasternal short/long) and mitral regurgitation assessment (apical four-chamber color Doppler, parasternal short/long color Doppler).

**Conclusions:** EchoFocus-Measure delivers rapid and reliable automated echo measurements across ages and lesions within diverse internal and real-world external cohorts, serving as a step toward scalable, global access to high-quality pediatric cardiovascular care.

## INTRODUCTION

Transthoracic echocardiography (echo) is a noninvasive, portable imaging modality that forms the foundation of pediatric cardiology worldwide by enabling diagnosis and longitudinal assessment of pediatric and congenital heart disease (CHD).^1^ Accurate measurements of ventricular function,^2^ chamber dimensions,^3^ and valvar structure and performance^4,5^ are essential for timely diagnosis, monitoring disease progression, and guiding clinical decision-making.

Reliable echo assessment requires specialized training that remains maldistributed globally. In many rural regions and low- and middle-income countries (LMICs), limited access to expert interpretation constrains the clinical utility of echo,^6–10^ contributing to delayed disease recognition and widening disparities in pediatric cardiovascular outcomes.^9,11^ Even in high-resource healthcare settings, substantial inter-operator and inter-institutional variability underscores the need for standardized, reproducible echo measurements to ensure consistent care.^12,13^

Recent advances in artificial intelligence (AI) have begun to reshape echo, particularly in adult populations where AI-based systems have progressed from automating individual measurements to supporting comprehensive, study-level interpretation.^14–19^ However, this progress lags in pediatric and CHD echo due to the unique challenges of anatomic variation and evolving physiologic states from infancy to adulthood. Consequently, prior efforts have largely focused on narrowly scoped tasks (e.g., recognizing standard imaging views, extracting individual measurements, or identifying specific CHD lesions),^12,13,20–23^ leaving an unmet need for broad, scalable, study-level measurement tools to support routine pediatric cardiac care.

To address this gap, we developed EchoFocus-Measure, a multi-task, study-level AI-echo model designed to generate 18 quantitative and 10 qualitative measurements. We evaluated real-world performance and generalizability using echos from 37 countries across five continents, with the goal of enabling scalable automated measurements for global standardization and expanding access to high-quality pediatric echo assessment worldwide.

## METHODS

This study is reported in accordance with the TRIPOD+AI 2024 guidelines.^24^

### Patient Population and Patient Assignment

Echo data were retrospectively obtained at Boston Children’s Hospital (BCH) from July 2015 through July 2025. All available transthoracic echos acquired during this period were screened for inclusion. Studies failing predefined quality control standards (described in “Data Retrieval, Pre-Processing, and Quality Control”) were excluded, yielding the main study cohort.

Echos were grouped based on acquisition site into internal studies (performed at BCH or affiliated outpatient centers) and external referral studies. The internal dataset was further subdivided by randomly allocating patients 80:20 to model development and testing sets, respectively. There was no patient overlap between development and test cohorts.

### Definition of Outcomes

For both internal and external cohorts, the ground truth outcome label was derived from the final clinical report generated by a BCH attending pediatric cardiologist with subspecialty training in noninvasive pediatric cardiac imaging. This approach holds the interpretation standard constant, allowing the external validation to primarily assess model robustness to image acquisition heterogeneity arising from different scanners, ultrasound systems, and imaging protocols.

The outcomes of interest were 18 quantitative measures and 10 qualitative measures. The 18 quantitative measures included the following: aortic valve diameter, aortic root diameter, left ventricular ejection fraction (LVEF), LV end-diastolic volume, LV posterior wall thickness, septal wall thickness, LV mass, LV end-systolic volume, mitral valve diameter, main pulmonary artery diameter, pulmonary valve diameter, tricuspid valve diameter, left atrial volume, right atrial volume, right ventricular longitudinal strain, LV circumferential strain, LV longitudinal strain, and RV free wall strain. All internal and external measurements were stored in a structured database within a custom hospital software. All measurements were transformed into unitless variables via z-score normalization or Box-Cox transformation. Model outcomes were evaluated using raw measurement values. The primary outcome was LVEF, which institutionally is measured via the bullet method (5/6 area-length method).^25^

The 10 qualitative measures were defined as at least moderate severity of the following: left ventricular outflow tract obstruction, aortic regurgitation, aortic stenosis, mitral regurgitation, right ventricular hypertension, tricuspid regurgitation, LV systolic dysfunction, pulmonary regurgitation, LV hypertrophy, and right ventricular outflow tract obstruction. To create these labels, we leveraged our institutional Fyler coding system—a detailed, decades-established anatomic classification system used at BCH and specifically designed for pediatric and CHD.^26^ For every echo, expert interpreting cardiac imagers assign Fyler codes that capture qualitative severity grading of ventricular and valvar function, in addition to structural cardiac lesions with high anatomic granularity. Each of the qualitative labels were marked as negative if they were qualified as less than moderate severity. Due to the natural history of CHD (e.g., tetralogy of Fallot) and institutional practice of Fyler code use, right ventricular outflow tract obstruction was grouped as a composite of pulmonary stenosis, right ventricle-to-pulmonary artery conduit stenosis, or right ventricular outflow tract obstruction.

### Data Retrieval, Pre-Processing, and Quality Control

Echo studies were obtained from the BCH picture archiving and communication system. Studies with fewer than 10 DICOM files were excluded. The remaining studies were then processed using a standardized pipeline adapted from PanEcho.^14^ For each study, raw two-dimensional echo videos were extracted directly from DICOM files. All data underwent thorough deidentification prior to analysis. Each frame was binarized to delineate the primary imaging region, and pixels outside the largest detected contour were concealed. Videos were then cropped to the central imaging area, resized to 256 x 256 using bicubic interpolation, and further anonymized by masking peripheral regions that could contain identifying information.^14^ *EchoFocus-Measure Model Architecture*

EchoFocus-Measure is designed to convert a full set of echo video clips from a single study into a comprehensive set of quantitative or qualitative measurements. The model integrates information across all videos to emulate the approach of a skilled clinician, who synthesizes multiple views to generate accurate cardiac assessments. The framework builds on a PanEcho backbone,^14^ enhanced with an additional transformer layer to allow the model to focus^27^ on the most informative video clips and capture complex inter-video relationships (Figure 1B).

**Figure 1:**
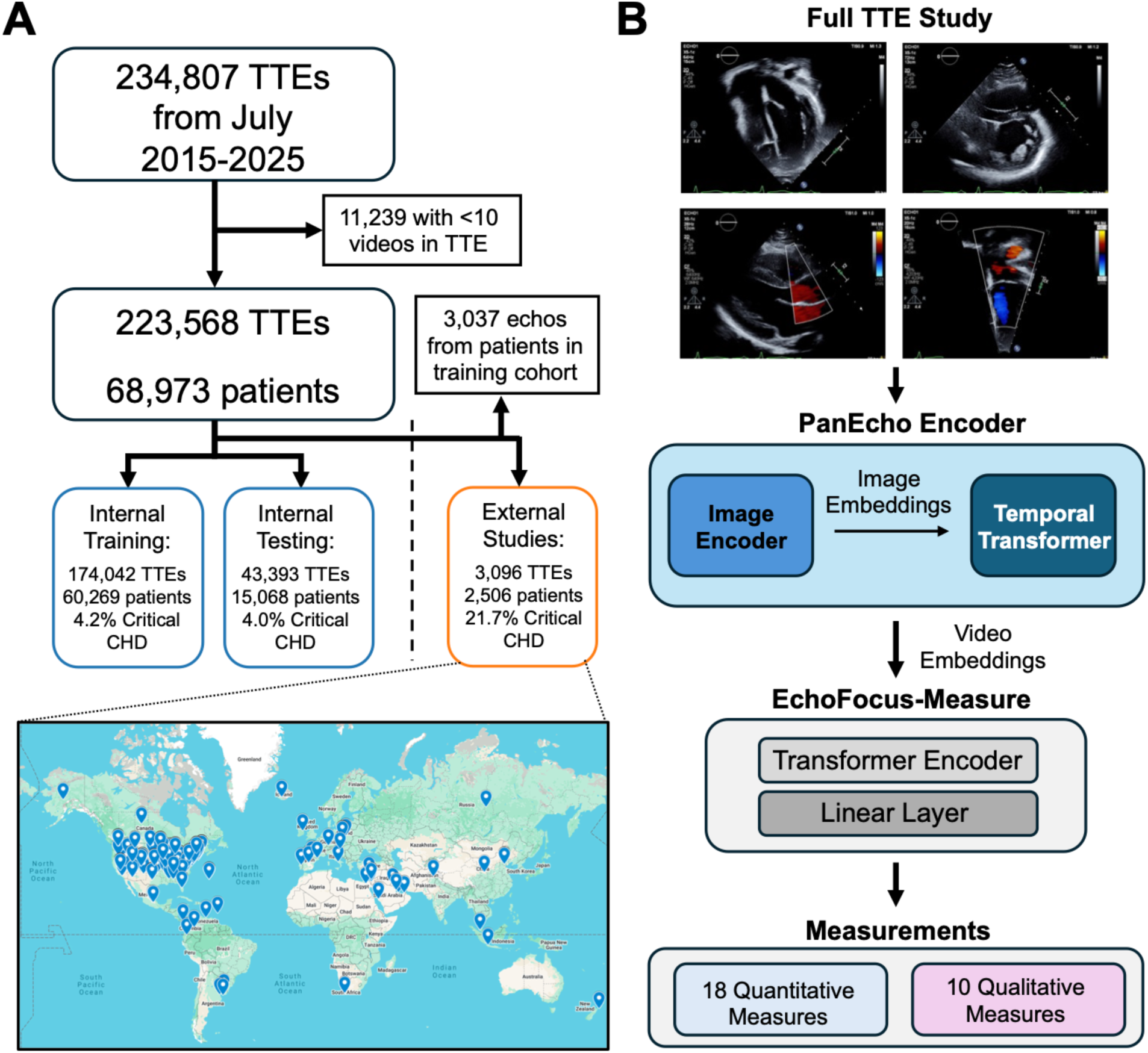
Study Design and Model Architecture. (**A**) Study design schematic with STROBE diagram showing initial patient selection and filtering to form the main cohort. Pins of origin countries for external patients inset. (**B**) EchoFocus-Measure architecture schematic and outcome targets. **Abbreviations:** transthoracic echo (TTE).

Each video is first converted into 16 random sets of 16 sequential frames (each called clips); each frame (image) is processed by a 2D convolutional neural network (ConvNeXt-T,^28^ pretrained on ImageNet) to extract rich feature representations. The resulting individual image embeddings are sequentially organized and passed through a temporal transformer with four layers and eight attention heads. Positional encodings preserve temporal order, and clip-level outputs are aggregated via mean pooling to generate a single clip embedding as a 768-dimensional vector.

EchoFocus-Measure then expands upon the original PanEcho architecture^14^ and incorporates an additional transformer that operates across all clip embeddings (number of videos x 16) to generate a single, study-level embedding. Unlike PanEcho, the transformer encoder analyzes all video embeddings collectively, enabling the model to identify patterns that emerge only when multiple views are considered together. The resulting study-level embedding is then passed through fully connected layers to produce predictions for measurements of interest.

Two separate models were trained: a regression model simultaneously predicting 18 quantitative echo measurements, and a binary classification model simultaneously predicting 10 qualitative measurements. For the regression model, echo studies were included if at least one quantitative measurement was available; missing measurement targets were masked and excluded from the loss computation.

### Model Training Strategy

For model development, echos from the internal development cohort were randomly divided at the patient level into training and validation subsets using an 80:20 split. The training subset was used to optimize model parameters, while the validation subset was reserved for model selection and early stopping. Pretrained PanEcho weights were frozen during training, enabling the optimization process to focus on the newly introduced study-level transformer encoder and downstream task-specific prediction layers.

Training was performed using the AdamW optimizer^29^ with a weight decay coefficient of 0.01. A dynamic learning rate schedule was employed, reducing the learning rate in response to plateaus in validation loss. Training stopped when no improvement in validation performance was observed for 10 consecutive epochs.

To enhance robustness to real-world echo variability, several regularization strategies were applied. Dropout^30^ was incorporated during training at a rate of 0.2 for weights and 0.5 for video clips to reduce overfitting and improve tolerance to incomplete or heterogeneous video inputs. In addition, consistent with prior PanEcho-based approaches,^14^ data augmentation techniques such as random cropping, rotation, and horizontal flipping were applied to mitigate sensitivity to image acquisition variability and noise.

Hyperparameter optimization was conducted through systematic exploration of key architectural and training parameters: the number of layers in the study-level transformer encoder (1, 5, 10, and 20); learning rates ranging from 1e-4 to 1e-2; and effective batch sizes between 32 and 128. The final classification model configuration was selected to minimize aggregate validation loss across all prediction tasks, while the final regression model was chosen to minimize LVEF validation error.

### Model Performance Assessment and Statistical Analysis

Regression model performance for continuous outcomes was evaluated using the median absolute error (MAE). Results are not reported for measurements with fewer than 10 available observations (e.g., strain from external echos) due to insufficient sample size. Where comparable endpoints existed, performance was benchmarked against the PanEcho framework.

For binary prediction tasks, discriminatory performance was evaluated using the area under the receiver operating characteristic curve (AUROC) and the area under the precision-recall curve. Results with less than 10 positive cases were not reported. To facilitate clinical interpretation, sensitivity, specificity, positive and negative likelihood ratios, positive and negative predictive values, and lift were calculated.

Operating thresholds for binary outcomes were selected based on the maximum Youden index, determined from the validation dataset and applied consistently across test cohorts. Unless otherwise specified, larger metric values reflect superior model performance. Statistical uncertainty was estimated via nonparametric bootstrapping with 1,000 resamples, and corresponding confidence intervals were reported for all primary performance measures.

### Subgroup Analysis

Subgroup analyses were performed on the test cohorts stratified by age groupings^31^ of age < 1 (infant), 1 ≤ age < 3, 3 ≤ age < 8, 8 ≤ age < 12, 12 ≤ age ≤ 18 years, and age > 18 years. Model discrimination within each age subgroup was assessed using AUROC. In addition, performance was assessed with subgroups of individual CHD lesions, as well as composites of critical and non-critical CHD (for definitions, see Supplementary Materials). Labels for CHD were generated using Fyler codes from the first BCH echo report per patient.

Performance was additionally evaluated in a cardiomyopathy subgroup, inclusive of dilated, hypertrophic, restrictive, arrhythmogenic, non-compaction, metabolic/mitochondrial, and unspecified cardiomyopathies. To account for the possibility that cardiomyopathy may not have been detected on the initial echo, patients were considered to have cardiomyopathy if they had a corresponding code on any echo in their record.

### Model Adjudication

To compare model versus human error, adjudication was performed. Four blinded sonographers remeasured 50 echo studies with discrepant LVEF measurements (MAE >50^th^ percentile), and 50 echo studies with discrepant aortic root measurements (MAE >50^th^ percentile). The latter was selected to focus on a valve measurement that carries clinical significance in the aortopathy population. Adjudicators were blinded to patient names, echo reports, model predictions, and to each other’s assessments.

Post-hoc analysis including calculating the human MAE (derived via leave-one-human-out methodology) and the EchoFocus-Measure MAE (derived using the median of blinded sonographer measurements as the ground truth). In addition, to assess agreement, the human-human (i.e., all four blinded sonographers) and the human-AI (i.e., all four blinded sonographers and AI) intraclass correlation coefficients were calculated.

### Model Interpretability

To enhance interpretability of model outputs, we conducted post-hoc attribution analyses using integrated gradients^32^ for predictions of LVEF and mitral regurgitation. For each task, we selected 25 echo studies with the smallest absolute prediction error (i.e., lowest MAE for LVEF; lowest error among studies positive for mitral regurgitation). Within each selected study, attribution scores were calculated to characterize the influence of individual video clips on the corresponding model prediction. Clips were ordered by attribution magnitude, and the ten most influential videos per study were retained for subsequent qualitative assessment.

The selected clips were then reviewed independently by a pediatric cardiology fellow, who identified and recorded the echo views represented among the model-prioritized inputs. *Data Availability and Software*

To support transparency and reproducibility, the EchoFocus-Measure model and associated source code are publicly accessible for non-commercial, academic research use at https://echofocus.org. Access to echo data derived from BCH is governed by institutional policies; requests will be evaluated to determine feasibility based on privacy, intellectual property, and regulatory considerations. When permitted, deidentified data and related materials will be shared under an institutional material transfer agreement for non-commercial, research purposes only. This study was conducted with approval from the BCH Institutional Review Board.

## RESULTS

### Patient Population Characteristics

There were 234,807 transthoracic echos at Boston Children’s Hospital meeting inclusion criteria. After excluding 11,239 echos with less than 10 DICOM files per study and 3,037 echos from overlapping patients in the training and external cohorts, there were 220,531 studies remaining, forming the main cohort (Figure 1A). Of those, 217,435 echos (64,403 patients) were from the internal cohort, and 3,096 echos (2,506 patients) were from the external cohort. The external international patients resided in 37 countries spanning five continents: North America, South America, Europe, Asia, and Africa.

As shown in Table 1, there were several differences between the internal (n=60,269 patients for model development; n=15,068 for testing) and external (n=2,506) cohorts. In general, the external cohort was more complex with higher rates critical CHD (21.7% versus 4.0-4.2%), non-critical CHD (33.0% versus 16.9-17.2%), and any CHD (43.8% versus 18.2-18.5%). Prevalence for each individual lesion is shown in Table 1. Cardiomyopathy was present in 1.8%, 1.5%, and 1.6% of patients in the internal development, internal test, and external test cohorts, respectively.

**Table 1:** Baseline Characteristics of Internal and External Cohorts.

|  | Internal Development | Internal Test | External |
| --- | --- | --- | --- |
| Patients | 60,269 | 15,068 | 2,506 |
| Sex (Male) | 32,162 (53.36%) | 8051 (53.43%) | 1,391 (55.51%) |
| ASD | 3,019 (5.01%) | 789 (5.24%) | 282 (11.25%) |
| Anomalous Coronaries | 299 (0.50%) | 72 (0.48%) | 33 (1.32%) |
| Bicuspid Aortic Valve | 1,213 (2.01%) | 286 (1.90%) | 123 (4.91%) |
| Double Aortic Arch | 82 (0.14%) | 17 (0.11%) | 6 (0.24%) |
| DORV | 156 (0.26%) | 42 (0.28%) | 79 (3.15%) |
| D-loop TGA | 228 (0.38%) | 69 (0.46%) | 44 (1.76%) |
| Ebstein Anomaly | 91 (0.15%) | 17 (0.11%) | 40 (1.60%) |
| HLHS | 188 (0.31%) | 48 (0.32%) | 63 (2.51%) |
| IAA | 36 (0.06%) | 13 (0.09%) | 8 (0.32%) |
| L-loop TGA | 65 (0.11%) | 15 (0.10%) | 48 (1.92%) |
| PAPVC | 262 (0.43%) | 67 (0.44%) | 24 (0.96%) |
| Patent Ductus Arteriosus | 4,649 (7.71%) | 1,110 (7.37%) | 179 (7.14%) |
| Right Aortic Arch | 464 (0.77%) | 117 (0.78%) | 38 (1.52%) |
| Tricuspid Atresia | 69 (0.11%) | 26 (0.17%) | 11 (0.44%) |
| Truncus arteriosus | 48 (0.08%) | 13 (0.09%) | 15 (0.60%) |
| Single Ventricle Disease | 313 (0.52%) | 87 (0.58%) | 93 (3.71%) |
| Tetralogy of Fallot | 505 (0.84%) | 114 (0.76%) | 56 (2.23%) |
| AVCD | 414 (0.69%) | 101 (0.67%) | 146 (5.83%) |
| VSD | 3,204 (5.32%) | 767 (5.09%) | 293 (11.69%) |
| Coarctation of the Aorta | 830 (1.38%) | 179 (1.19%) | 82 (3.27%) |
| Pulmonary Atresia | 255 (0.42%) | 44 (0.29%) | 56 (2.23%) |
| TAPVC | 59 (0.10%) | 25 (0.17%) | 17 (0.68%) |
| Any Non-Critical CHD | 10,346 (17.17%) | 2,550 (16.92%) | 826 (32.96%) |
| Any Critical CHD | 2,547 (4.23%) | 606 (4.02%) | 543 (21.67%) |
| Any CHD | 11,153 (18.51%) | 2,741 (18.19%) | 1,097 (43.77%) |
| Any Cardiomyopathy | 1,059 (1.76%) | 227 (1.51%) | 39 (1.56%) |

Data presented as median (interquartile range).
**Abbreviations:** atrial septal defect (ASD); double outlet right ventricle (DORV); transposition of the great arteries (TGA); hypoplastic left heart syndrome (HLHS); interrupted aortic arch (IAA); partial anomalous pulmonary venous connection (PAPVC); atrioventricular canal defect (AVCD); ventricular septal defect (VSD); total anomalous pulmonary venous connection (TAPVC).

As shown in Table 2, there were accompanying differences in echo characteristics between the internal (n=174,042 echos for model development; n=43,393 echos for testing) and external patients (n=3,096 echos). The internal model development and testing cohorts had 9.1 million and 2.3 million echo videos respectively, totaling >11 million. There were 47 [IQR, 36-66] videos per study for the internal model development and test cohort. The external cohort had 289,613 videos, with 44 [IQR 29-62] videos per study. The external cohort was younger (median age at echo 3.5 [IQR 0.6-11.1] years) compared to the internal development (median age at echo 8.5 [IQR 1.1-16.6] years) and test (median age at echo 8.5 [1.2-16.5] years) cohorts. Accordingly, the external cohort had smaller raw LV end-diastolic volumes, LV masses, LV end-systolic volumes, valvar measurements, and atrial volumes.

**Table 2:** Echo Characteristics of Internal and External Cohorts.

|  | Internal Development | Internal Test | External |
| --- | --- | --- | --- |
| Number of Echos | 174,042 | 43,393 | 3,096 |
| Videos | 9,099,124 | 2,271,924 | 289,613 |
| Videos Per Study | 47 (36, 66) | 47 (36, 66) | 44 (29, 62) |
| Age at Echo | 8.49 (1.13,16.57) | 8.46 (1.18,16.46) | 3.52 (0.55,11.11) |
| Echos with $\geq 1$ Measure | 150,836 | 37,652 | 1,915 |
| Aortic Valve Diameter (cm) | 1.66 (1.14,2.00) | 1.66 (1.15,1.99) | 1.35 (0.91,1.81) |
| Aortic Root Diameter (cm) | 2.37 (1.70,2.88) | 2.38 (1.72,2.88) | 2.00 (1.40,2.60) |
| LVEF (%) | 61 (58,65) | 61 (58,65) | 62 (57,66) |
| LVEDV (mL) | 73.20 (27.40,127.00) | 75.30 (27.30,127.10) | 61.45 (20.22,116.15) |
| LV Posterior Wall Thickness (cm) | 0.66 (0.52,0.78) | 0.66 (0.52,0.78) | 0.66 (0.50,0.82) |
| Septal Wall Thickness (cm) | 0.69 (0.55,0.83) | 0.69 (0.55,0.82) | 0.71 (0.57,0.93) |
| LV Mass (g) | 59.40 (23.50,105.70) | 60.90 (23.50,105.90) | 49.40 (19.45,98.20) |
| LVESV (mL) | 27.40 (10.40,48.80) | 28.10 (10.32,48.80) | 22.80 (7.60,43.75) |
| Mitral Valve Diameter (cm) | 2.09 (1.29,2.61) | 2.10 (1.28,2.62) | 1.77 (1.26,2.42) |
| MPA Diameter (cm) | 1.85 (1.10,2.37) | 1.87 (1.12,2.39) | 1.50 (1.02,2.16) |
| Pulmonary Valve Diameter (cm) | 1.67 (0.91,2.36) | 1.63 (0.92,2.35) | 1.22 (0.91,1.81) |
| Tricuspid Valve Diameter | 2.20 (1.51,2.70) | 2.20 (1.51,2.70) | 2.00 (1.40,2.59) |
| Left Atrial Volume (mL) | 37.60 (23.80,53.50) | 37.80 (23.80,53.90) | 28.90 (14.70,42.90) |
| Right Atrial Volume (mL) | 27.90 (13.60,48.00) | 29.90 (13.60,49.00) | 24.60 (17.60,43.40) |
| RV Longitudinal Strain (%) | -20.50 (-24.00,-16.50) | -20.50 (-24.10,-16.70) | — |
| LV Circumferential Strain (%) | -28.90 (-32.10,-25.20) | -29.00 (-32.30,-25.20) | — |
| LV Longitudinal Strain (%) | -20.80 (-23.20,-18.10) | -20.80 (-23.20,-18.10) | — |
| RV Free Wall Strain (%) | -22.50 (-27.50,-17.90) | -22.80 (-27.60,-18.00) | — |
| LVOTO | 1,666 (0.96%) | 337 (0.78%) | 89 (2.87%) |
| Aortic Regurgitation | 3,066 (1.76%) | 816 (1.88%) | 135 (4.36%) |
| Aortic Stenosis | 1,707 (0.98%) | 306 (0.71%) | 37 (1.20%) |
| Mitral Regurgitation | 5,395 (3.10%) | 1,368 (3.15%) | 227 (7.33%) |
| RV Hypertension | 17,195 (9.88%) | 4,300 (9.91%) | 256 (8.27%) |
| Tricuspid Regurgitation | 9,215 (5.29%) | 2,129 (4.91%) | 220 (7.11%) |
| LV Systolic Dysfunction | 7,491 (4.30%) | 1,831 (4.22%) | 58 (1.87%) |
| Pulmonary Regurgitation | 5,339 (3.07%) | 1,382 (3.18%) | 105 (3.39%) |
| LV Hypertrophy | 2,614 (1.50%) | 608 (1.40%) | 62 (2.00%) |
| RVOTO | 5,065 (2.91%) | 1,242 (2.86%) | 172 (5.56%) |

Data presented as median (interquartile range).
**Abbreviations:** left ventricular end-diastolic volume (LVEDV); left ventricular end-systolic volume (LVESV); main pulmonary artery (MPA); right ventricle (RV); LV outflow tract obstruction (LVOTO); RV outflow tract obstruction (RVOTO).

The higher rates of external CHD were accompanied by higher rates of qualitative outcomes such as biventricular outflow tract obstruction, valvar regurgitation, aortic stenosis (1.2% versus 0.7-1.0%), and LV hypertrophy (2.0% versus 1.4-1.5%). The internal cohort had higher rates of LV systolic dysfunction and right ventricular hypertension (Table 2).

### EchoFocus-Measure Regression Performance

Regression model performance of EchoFocus-Measure for 18 individual measurements during internal and external testing is shown in Figure 2. Internally, MAE was: 0.09 cm (aortic valve diameter), 0.13 cm (aortic root diameter), 2.8% (LVEF), 5.8 mL (LV end-diastolic volume), 0.05 cm (LV posterior wall thickness), 0.05 cm (septal wall thickness), 5.1 g (LV mass), 2.5 mL (2LV end-systolic volume), 0.14 cm (mitral valve diameter), 0.17 cm (main pulmonary artery diameter), 0.14 cm (pulmonary valve diameter), 0.18 cm (tricuspid valve diameter), 4.8 mL (left atrial volume), 5.5 mL (right atrial volume), 2.6% (right ventricular longitudinal strain), 2.6% (LV circumferential strain), 1.9% (LV longitudinal strain), and 3.2% (right ventricular free wall strain). During external validation, there was variable increase in MAE among metrics; for example, external LVEF MAE increased to 3.8%, LV end-diastolic volume MAE increased to 13.8 mL, and aortic root diameter MAE increased to 0.27 cm.

**Figure 2:**
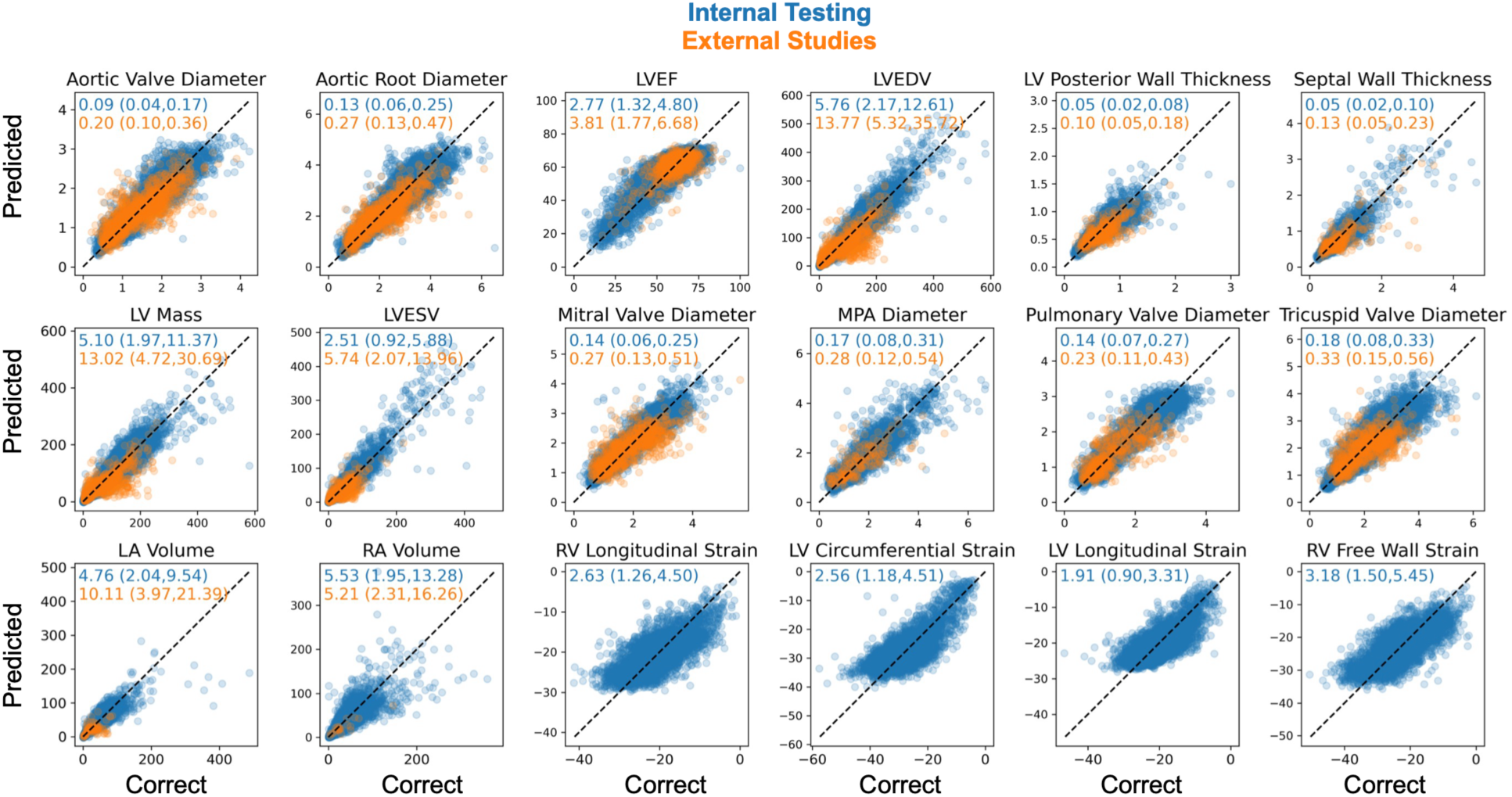
EchoFocus-Measure Regression Task Performance. Internal (blue) and external (orange) performance of EchoFocus-Measure to predict 18 measurements in the pediatric and CHD population. MAE values inset. Dotted line represents the identity line. **Abbreviations:** left ventricular ejection fraction (LVEF); LV end-diastolic volume (LVEDV); LV end-systolic volume (LVESV); main pulmonary artery (MPA); left atrium (LA); right atrium (RA); right ventricle (RV).

During benchmarking, EchoFocus-Measure outperformed PanEcho in predicting LVEF in the overall internal (MAE 2.8% versus 7.3%) and external (MAE 3.8% versus 7.9%) cohorts and in specific age and lesion subgroups (Table 3). EchoFocus-Measure performance remained similar for ages >3, followed by a slight drop for ages 1-3 (internal MAE 3.0%, external MAE 3.8%) and age <1 (internal MAE 3.2%, external MAE 4.4%). PanEcho performance started to drop for ages <8 years old.

**Table 3:** Benchmarking EchoFocus-Measure to PanEcho.

|  | Internal |  | External |  |
| --- | --- | --- | --- | --- |
|  | EchoFocus-Measure | PanEcho | EchoFocus-Measure | PanEcho |
| Overall Cohort | 2.77 (1.32,4.80) | 7.30 (4.04,10.97) | 3.81 (1.77,6.68) | 7.86 (3.95,12.34) |
| **Age Subgroups (years)** |  |  |  |  |
| <1 | 3.18 (1.50,5.60) | 7.54 (4.08,11.52) | 4.43 (2.01,7.70) | 8.20 (4.15,12.59) |
| 1-3 | 2.96 (1.35,5.06) | 8.43 (4.70,12.16) | 3.81 (2.04,6.82) | 9.16 (4.42,14.32) |
| 3-8 | 2.60 (1.25,4.50) | 8.08 (5.05,11.40) | 3.96 (2.08,6.34) | 8.51 (4.63,12.66) |
| 8-12 | 2.73 (1.32,4.58) | 7.35 (4.13,10.99) | 3.41 (1.66,6.55) | 7.67 (3.80,12.30) |
| 12-18 | 2.57 (1.23,4.36) | 6.39 (3.39,9.84) | 3.29 (1.52,5.19) | 6.57 (2.89,11.01) |
| ≥18 | 2.75 (1.33,4.76) | 7.04 (3.77,10.63) | 3.38 (1.50,7.71) | 5.61 (3.19,10.55) |
| **Lesion Subgroups** |  |  |  |  |
| AP window | 2.55 (0.77,4.93) | 11.51 (10.11,14.10) | — | — |
| ASD | 3.14 (1.48,5.59) | 8.18 (4.69,12.34) | 4.62 (2.07,6.78) | 8.49 (5.39,12.94) |
| Anomalous Coronaries | 2.75 (1.48,4.44) | 6.30 (2.97,10.02) | — | — |
| Bicuspid Aortic Valve | 3.04 (1.41,5.40) | 8.13 (4.86,12.29) | 5.23 (2.38,7.73) | 7.93 (3.36,12.96) |
| Cor Triatriatum | 3.10 (1.71,5.69) | 11.73 (8.92,16.34) | — | — |
| Double Aortic Arch | 1.78 (0.65,3.08) | 7.98 (5.18,8.84) | — | — |
| DORV | 4.17 (2.07,8.04) | 9.00 (5.32,12.85) | — | — |
| D-loop TGA | 2.36 (1.06,4.13) | 9.06 (6.16,11.45) | — | — |
| Ebstein Anomaly | 3.78 (1.63,6.92) | 8.87 (3.54,11.66) | — | — |
| HLHS | 5.76 (2.81,8.25) | 12.98 (7.12,15.57) | — | — |
| Interrupted Aortic Arch | 3.46 (1.32,5.63) | 10.63 (6.41,14.32) | — | — |
| L-loop TGA | 3.95 (1.30,7.19) | 13.12 (7.03,15.84) | — | — |
| PAPVC | 3.00 (1.43,4.78) | 9.56 (5.90,13.53) | — | — |
| PDA | 3.13 (1.49,5.63) | 8.31 (4.66,12.30) | 5.29 (2.52,8.42) | 8.16 (4.88,13.39) |
| Right Aortic Arch | 3.04 (1.52,5.26) | 7.67 (4.60,11.52) | — | — |
| Tricuspid atresia | 3.18 (1.97,5.84) | 6.74 (3.62,10.32) | — | — |
| Truncus Arteriosus | 2.46 (1.58,4.51) | 7.63 (5.16,10.57) | — | — |
| Any SV Disease | 4.84 (2.50,8.07) | 9.94 (5.55,14.89) | — | — |
| Tetralogy of Fallot | 3.08 (1.50,5.35) | 9.08 (5.68,12.92) | — | — |
| AVCD | 3.42 (1.65,6.21) | 8.03 (4.48,12.02) | 4.77 (1.93,8.77) | 9.41 (6.41,15.60) |
| VSD | 3.11 (1.43,5.44) | 7.89 (4.48,11.61) | 3.89 (1.67,6.71) | 7.93 (4.23,11.31) |
| Coarctation of the Aorta | 3.48 (1.53,6.38) | 8.70 (4.71,12.82) | 5.77 (3.49,8.50) | 8.06 (5.02,11.96) |
| Pulmonary Atresia | 3.26 (1.50,6.35) | 8.34 (5.21,12.41) | — | — |
| TAPVC | 3.20 (1.83,4.21) | 5.16 (1.02,9.00) | — | — |
| Critical Aortic Stenosis | 3.33 (2.02,6.80) | 12.23 (8.03,15.04) | — | — |
| Critical Pulmonary Stenosis | 2.24 (1.33,6.52) | 5.63 (3.29,9.34) | — | — |
| Any Non-Critical CHD | 2.98 (1.42,5.32) | 8.02 (4.60,11.90) | 4.29 (1.99,7.27) | 7.76 (4.36,12.07) |
| Any Critical CHD | 3.32 (1.55,6.08) | 8.72 (5.14,12.76) | 5.48 (2.46,8.87) | 8.69 (4.83,13.26) |
| Any CHD | 2.98 (1.41,5.31) | 8.07 (4.66,11.93) | 4.36 (1.96,7.35) | 7.89 (4.38,12.30) |
| No CHD | 2.60 (1.25,4.39) | 6.73 (3.63,10.05) | 3.33 (1.59,6.10) | 7.81 (3.43,12.18) |
| Any Cardiomyopathy | 3.23 (1.62,5.64) | 8.28 (4.35,12.83) | 5.07 (2.00,8.09) | 10.51 (4.72,15.75) |

**Abbreviations:** aortopulmonary (AP); atrial septal defect (ASD); double outlet right ventricle (DORV); transposition of the great arteries (TGA); hypoplastic left heart syndrome (HLHS); partial anomalous pulmonary venous connection (PAPVC); patent ductus arteriosus (PDA); single ventricle (SV); atrioventricular canal defect (AVCD); ventricular septal defect (VSD); total anomalous pulmonary venous connection (TAPVC); congenital heart disease (CHD).

EchoFocus-Measure LVEF performance was relatively stable across all CHD lesions (internal MAE <5% except hypoplastic left heart syndrome). Similar trends were noted externally, with a maximal MAE of 5.8%. Interestingly, for L-loop transposition of the great arteries, MAE was 4.0% internally for EchoFocus-Measure, compared to 13.1% for PanEcho.

Among non-critical CHD studies, EchoFocus-Measure exhibited a LVEF MAE of 3.0% internally and 4.3% externally. LVEF MAE increased slightly for critical CHD (3.3% internal, 5.5% external) and decreased for patients without CHD (2.6% internal, 3.3% external). In the cardiomyopathy subgroup, LVEF MAE was 3.2% internally and 5.1% externally. PanEcho showed similar trends but with higher errors; for example, LVEF MAE for critical CHD was 8.7% both internally and externally. As illustrated in Figure S1, EchoFocus-Measure outperformed PanEcho for six additional measurements: LV end-diastolic volume, LV end-systolic volume, septal wall thickness, LV posterior wall thickness, left atrial volume, and aortic root diameter.

### Sonographer Adjudication

Discrepant internal test cases (>50^th^ percentile MAE) for LVEF and aortic root diameter were reviewed by four experienced blinded sonographers.

As shown in Figure 3A, for LVEF EchoFocus-Measure clustered with the original measurement and sonographers 1, 2, 4, whereas sonographer 3 did not. When compared to median sonographer LVEF measurements, the adjudicated EchoFocus-Measure MAE of 2.4% was within the sonographer MAE of 3.7%. The inter-sonographer intraclass correlation coefficient was 0.47; when adding EchoFocus-Measure to the pool of sonographer measurements, the intraclass correlation coefficient was unchanged (0.47).

**Figure 3.**
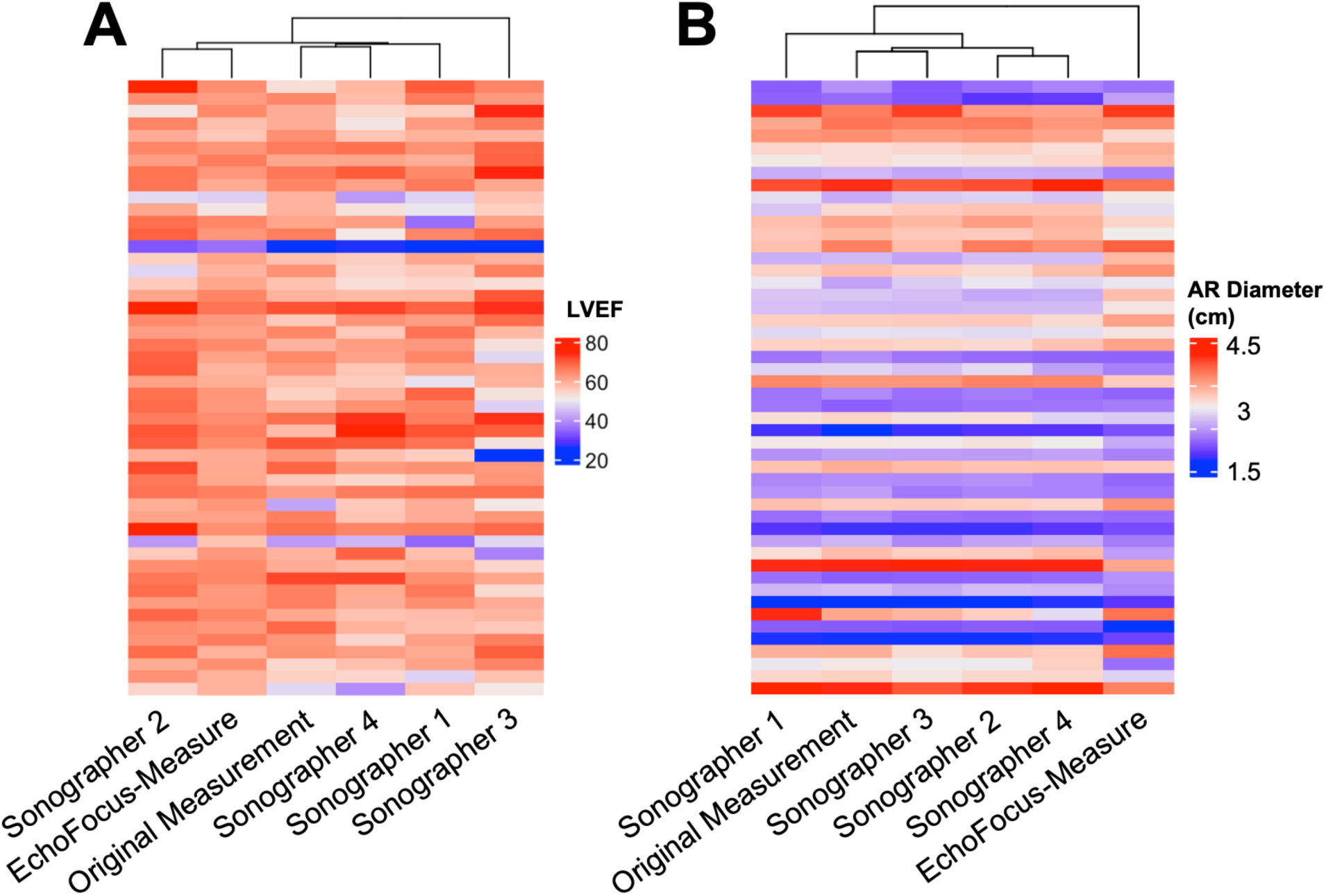
Sonographer Adjudication of Discrepant Cases. Expert adjudication was performed on 50 discrepant (**A**) LVEF cases and (**B**) 50 discrepant aortic root diameter cases. Heatmap with hierarchical clustering displaying measurements of individual sonographers versus EchoFocus-Measure versus the original measurement.

**Figure 4:**
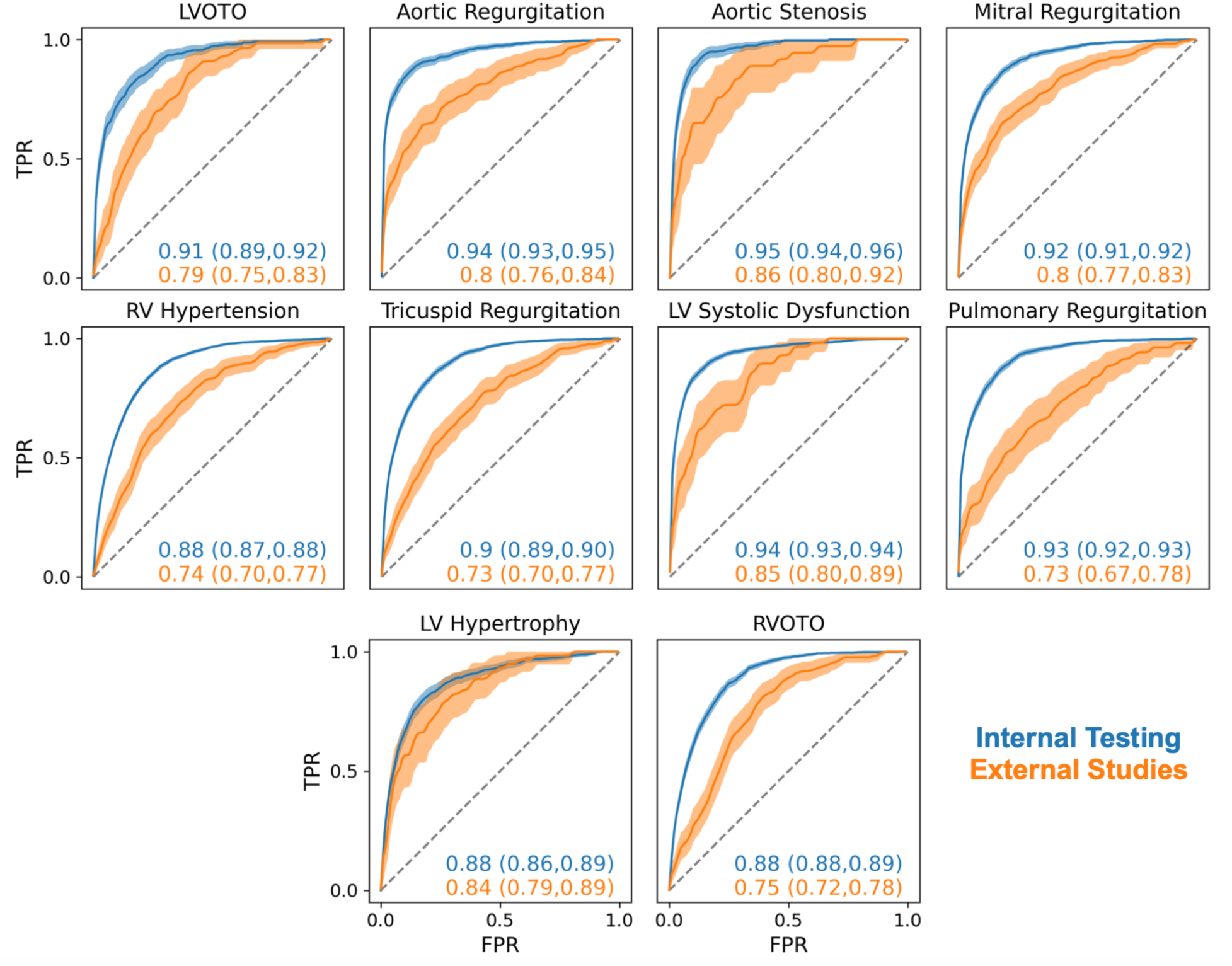
EchoFocus-Measure Performance for Qualitative Outcomes. Performance of the qualitative EchoFocus-Measure model to predict 10 qualitative measures during internal (blue) and external (orange) testing. Dotted line represents chance. 95% confidence intervals are shown using bootstrapping. **Abbreviations:** Left ventricular outflow tract obstruction (LVOTO); right ventricle (RV); RV outflow tract obstruction (RVOTO).

As shown in Figure 3B, EchoFocus-Measure clustered alone when estimating aortic root diameter. When compared to median sonographer aortic root diameter measurements, the adjudicated EchoFocus-Measure MAE of 0.53 cm was outside the sonographer MAE of 0.05 cm. The near perfect inter-sonographer intraclass correlation coefficient was 0.96; when adding EchoFocus-Measure to the pool of measurements, the intraclass correlation coefficient dropped to 0.91.

### EchoFocus-Measure Qualitative Outcome Performance

EchoFocus-Measure performance to detect 10 qualitative outcomes is shown in Figure 3. Internally, performance ranged from AUROC 0.88 (at least moderate RV hypertension, LV hypertrophy, and RV outflow tract obstruction) to 0.95 (at least moderate aortic stenosis). During external testing, there was a modest drop in performance, with AUROC ranging from 0.73 (pulmonary and tricuspid regurgitation) to 0.86 (aortic stenosis). Individual performance metrics for internal and external testing are shown in Tables S1 and S2.

For two valvar measurements of interest (aortic and mitral regurgitation), subgroup analysis was performed. As shown in Table S3, internal performance was highest for ages 3-18. No clear trend was apparent for external studies.

### Model Explainability

In model explainability analyses (Figure 5), EchoFocus-Measure assigned the highest attention for LVEF assessment to the apical four-chamber, parasternal long-axis, and parasternal short-axis views. For mitral regurgitation, the model similarly prioritized these same views, with Color Doppler clips being preferentially selected in most cases.

**Figure 5:**
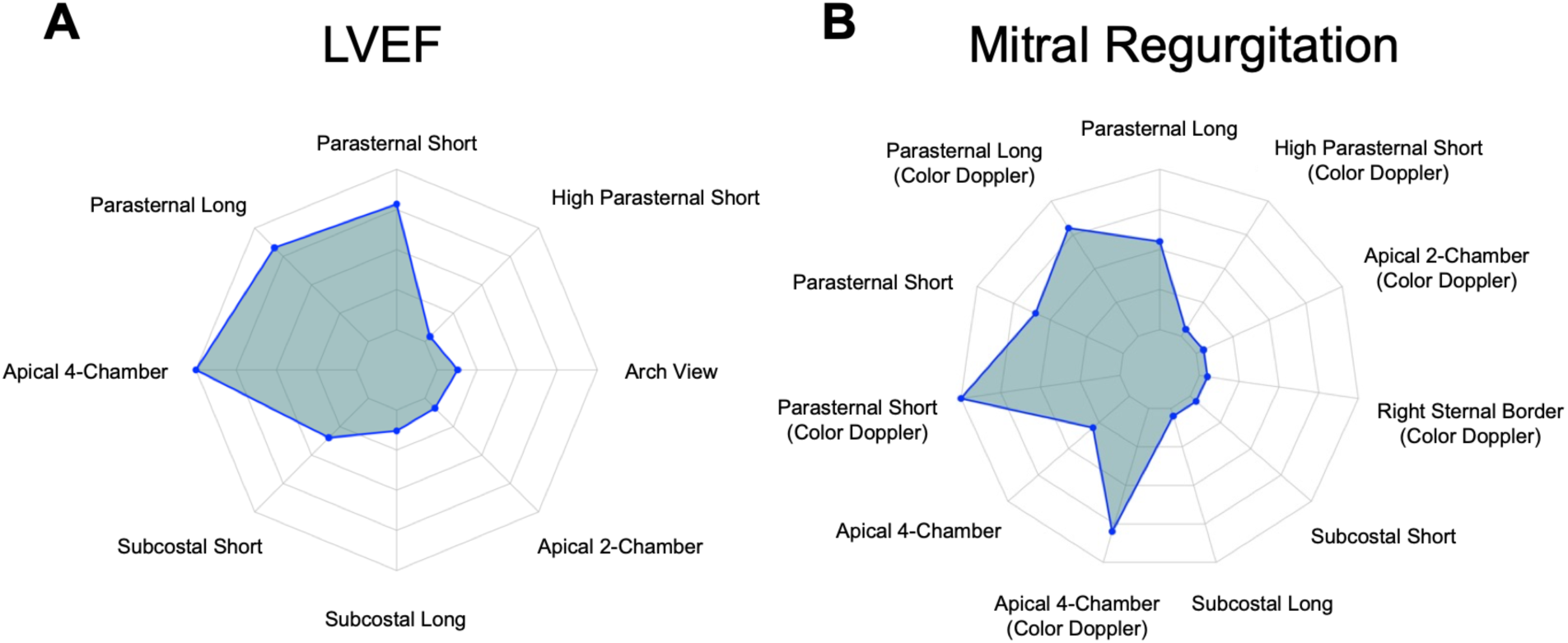
Model Explainability Analysis. Radar plots of top views selected during explainability analysis for (**A**) LVEF and (**B**) mitral regurgitation.

## DISCUSSION

EchoFocus-Measure is the first study-level, multi-task AI echo platform designed for automated evaluation of a broad set of common pediatric echo measurements, encompassing 18 quantitative and 10 qualitative parameters. The method extends the PanEcho framework with a clinically inspired study-level attention module that prioritizes diagnostically informative views in a manner analogous to expert cardiac imagers. By leveraging the largest pediatric and CHD echo dataset reported to date (>11 million videos), we address longstanding limitations related to sample size and age- and lesion-level heterogeneity that have constrained pediatric AI-echo development. We demonstrate that EchoFocus-Measure achieves expert-level performance for key functional measurements (including LVEF) across age, CHD lesion, and cardiomyopathy subgroups. Qualitative measurements showed strong internal performance with a modest decline on external testing. Notably, external performance herein was comparable to the internally reported performance of prior pediatric AI-echo studies focused on individual measurements, including mitral regurgitation (AUROC 0.75-0.84)^23^ and LVEF (MAE 3.7%).^12^ Finally, model explainability analyses suggest EchoFocus-Measure prioritizes clinically expected views for LVEF and mitral regurgitation prediction, lending trust to clinicians. Collectively, these findings suggest that EchoFocus-Measure is a step towards broadly delivering quality pediatric echo assessments across diverse clinical settings, with the potential to expand access to care, accelerate patient triage, standardize measurements, and streamline sonographer workflows.

### Clinical Need for Automated Echo Measurements

Pediatric heart failure is an underrecognized and growing global health challenge.^33–35^ In 2021, an estimated 6 million children were affected worldwide, a number forecasted to sharply rise by 2050.^33–35^ These figures likely underestimate the true burden because pediatric heart failure is systematically underdiagnosed in LMICs.^33^ Furthermore, the leading causes of pediatric heart failure^33^—CHD (48%), cardiomyopathy (20%), and rheumatic heart disease (11%)—are also major drivers of heart failure in adulthood; for example, the rapidly growing adult CHD population is at nearly 10-fold greater risk of heart failure,^36^ which represents the leading cause of mortality in this population.^37^ Rheumatic heart disease alone, which disproportionately affects children and young adults in resource-limited settings, impacts an estimated 40 million individuals globally and accounts for nearly 300,000 deaths annually.^38^

Across these conditions, delayed recognition of ventricular dysfunction and valvular disease leads to worse clinical trajectories for children and adults who might otherwise benefit from early referral, timely intervention, and guideline-directed therapy. Scalable and accessible technologies are urgently needed to facilitate earlier disease detection and enable clinically actionable decision-making across diverse care environments. This need is particularly acute in LMICs, where shortages of clinicians with specialized expertise in pediatric cardiology are profound,^9,10,39^ and where the burden of diseases such as rheumatic heart disease remains high. Even in well-resourced healthcare systems, substantial inter-operator and inter-institutional variability in echo measurements persists, underscoring the ongoing challenge and need for achieving standardized, reproducible quantitative assessment.^12,13^

### Clinical Implications of EchoFocus-Measure

With this context, EchoFocus-Measure was developed to benefit both low-resource and high-resource settings. In resource-limited settings, EchoFocus-Measure may support triage by identifying patients with ventricular dysfunction or clinically significant left-sided valvar disease, including patterns suggestive of rheumatic heart disease or congenital valvulopathies. LVEF error in external cohorts (MAE 3.8%) was comparable to human variability (MAE 3.7%), supporting the use of automated LVEF assessment for identifying patients at risk of systolic dysfunction. In addition, external left-sided valvar abnormality AUROCs of 0.80-0.86 and external positive predictive values of approximately 25% for mitral regurgitation suggest that the model can enrich for higher-risk patients who may benefit from earlier referral or further expert evaluation (Table S2).

Even in well-resourced healthcare systems, automated echo measurements offer important benefits. Substantial inter-operator and inter-institutional variability in measurements can adversely affect consistency and quality of care.^12,13^ Moreover, routine quantitative measurements consume significant clinician/sonographer time and could be reliably automated, allowing expert effort to be redirected toward higher-value interpretive and clinical decision-making tasks. EchoFocus-Measure achieves sonographer-level accuracy for LVEF measurement and demonstrates stable performance across a broad range of ages and CHD lesion types, supporting its use for automated measurements and quality assurance to streamline sonographer workflow. In the adult echo literature, a comparable AI-based system has been shown to be non-inferior to sonographers for LVEF assessment in a blinded, randomized trial, while also reducing interpretation time for both sonographers and cardiologists.^40^ EchoFocus-Measure could fill a similar role in pediatric practice, although prospective evaluation will be required to rigorously assess its impact on workflow and clinical decision-making.

### Importance of Real-World Deployment

A key finding of this study was the decline in model performance observed in a large, geographically and demographically distinct external cohort, reflecting conditions expected during deployment across diverse real-world care environments. This attenuation in performance is not unexpected given the greater clinical complexity, younger patient age distribution, and substantial heterogeneity in echo acquisition and processing across institutions. Variability in vendor-specific image pipelines, operator-dependent acquisition techniques, image quality, and local imaging protocols introduces domain shifts that can meaningfully affect model performance when models are applied beyond their development settings.

These results highlight the importance of diverse cohort training, rigorous external validation, and ongoing performance monitoring as AI-based echo tools are deployed across heterogeneous health systems globally. Strategies such as retraining with more diverse data, domain-aware calibration, and continual evaluation across regions and care contexts will be critical to ensure reliable performance as such tools are extended to settings with differing resources, workflows, and patient populations.

### Limitations and Future Directions

Several limitations warrant consideration. First, although model performance for LVEF was within the range of human variability, accuracy for other quantitative measurements was more variable, indicating opportunities for further improvement. Future work will focus on enhancing performance and generalizability through multiple complementary strategies, including exploration of other backbone architectures (such as EchoPrime)^19^ or learning approaches (e.g., adversarial learning^41^), development of pediatric cardiology-specific foundation models to learn more robust and anatomically informed representations, and incorporation of multi-institutional or federated learning approaches to better capture heterogeneity across both large referral centers and smaller care settings. Second, despite broad geographic diversity in the external validation cohort, certain regions with substantial unmet clinical need—notably sub-Saharan Africa—were not represented. As a result, generalizability to settings with the greatest burden of pediatric heart disease and the most constrained access to specialty care cannot be assumed and will require targeted evaluation. Third, the current models rely on transthoracic echos acquired by trained sonographers. Extension to low-resource or point-of-care ultrasound environments will necessitate additional validation on portable imaging systems, where image quality and operator variability may differ substantially. Finally, while post-hoc explainability analyses using integrated gradients provided insight into model behavior, further work is needed to determine how such explanations influence clinician trust, interpretability, and decision-making in real-world settings.

Future directions should therefore prioritize continued model refinement with an emphasis on robustness to heterogeneous acquisition conditions, prospective multi-site evaluation across diverse healthcare environments, and formal assessment of clinical utility, workflow integration, and impact on patient triage and outcomes.

## Conclusions

EchoFocus-Measure demonstrates that large-scale, multi-task AI models can provide accurate, automated echo measurements in pediatric populations using routine transthoracic echo. The model outperformed the PanEcho benchmark and achieved external performance comparable to internally reported results in the existing pediatric AI-echo literature for selected measurements. At the same time, these findings underscore the critical importance of rigorous external validation as such tools are extended across heterogeneous care environments. By transparently characterizing both strengths and limitations, this work establishes a foundation for prospective evaluation and iterative deployment strategies aimed at enabling equitable, scalable access to high-quality pediatric cardiac care worldwide.

## ACKNOWLEDGMENTS

The authors would like to acknowledge Boston Children’s Hospital’s High-Performance Computing Resources Clusters Enkefalos 3 (E3) made available for conducting the research reported in this publication.

## SUPPLEMENTAL MATERIAL

Tables S1-S3

Figure S1

