## Supplementary Materials for "Deep Learning-Based Automated Echocardiographic Measurements in Pediatric and Congenital Heart Disease"

**Running Title:** AI for Automated Pediatric Echo Measurements

Platon Lukyanenko*,[a] Sunil Ghelani*,[b] Yuting Yang,[a] Bohan Jiang,[a] Timothy Miller,[a] Peter Higgins,[b] Manouk Kirakosian,[b] Kaitlyn Tracy,[b] Janet Kane,[b] David Harrild,[b] John Triedman,[b] Andrew J. Powell,[b] Tal Geva,[b] William G. La Cava**,[a], Joshua Mayourian**,[b]

[a] Computational Health Informatics Program, Boston Children's Hospital, Department of Pediatrics, Harvard Medical School, Boston, MA, USA
[b] Department of Cardiology, Boston Children's Hospital, Department of Pediatrics, Harvard Medical School, Boston, MA, USA

* Co-First Authors
** Co-Senior Authors

**Address for correspondence:**
Joshua Mayourian
Department of Cardiology, Boston Children's Hospital
300 Longwood Avenue
Boston, MA 02115


*CHD Definitions*

The 12 critical CHD lesions predicted were double outlet right ventricle, D-loop transposition of the great arteries, Ebstein anomaly, hypoplastic left heart syndrome, tricuspid atresia, truncus arteriosus, any single ventricle disease (defined as “single ventricle”, “single left ventricle”, “single right ventricle”, tricuspid atresia, and hypoplastic left heart syndrome), tetralogy of Fallot, atrioventricular canal defect, coarctation of the aorta, pulmonary atresia, and total anomalous pulmonary venous connection. The composite critical CHD outcome includes each of these individual lesions, in addition to anomalous left coronary artery from the pulmonary artery, aortopulmonary window, double outlet left ventricle, interrupted aortic arch, critical aortic stenosis, and critical pulmonary stenosis.

The 8 non-critical CHD lesions predicted were atrial septal defect, anomalous coronaries, bicuspid aortic valve, left superior vena cava, partial anomalous pulmonary venous connection, patent ductus arteriosus, right aortic arch, and ventricular septal defect. The composite non-critical CHD outcome includes each of these individual lesions, in addition tocor triatriatum, double aortic arch, [I,D,D] transposition of the great arteries, left pulmonary artery sling, [S,L,L] transposition of the great arteries, and vascular ring.

**Table S1: Internal Performance Metrics for Qualitative Outcomes**

| Outcome | N (%) | AUROC | AUPRC | Lift | PPV | NPV | Sensitivity | Specificity | LR+ | LR- |
|---|---|---|---|---|---|---|---|---|---|---|
| LVOTO | 337 (0.78%) | 0.91 (0.89,0.92) | 0.18 (0.14,0.22) | 23.06 (17.98,28.86) | 0.03 (0.02,0.03) | 1.0 (1.00,1.00) | 0.88 (0.84,0.91) | 0.75 (0.75,0.76) | 3.54 (3.39,3.70) | 0.16 (0.12,0.21) |
| Aortic Regurgitation | 816 (1.88%) | 0.94 (0.93,0.95) | 0.53 (0.49,0.57) | 28.23 (26.02,30.75) | 0.15 (0.14,0.16) | 1.0 (1.00,1.00) | 0.84 (0.81,0.86) | 0.91 (0.91,0.91) | 9.45 (9.03,9.87) | 0.18 (0.15,0.21) |
| Aortic Stenosis | 306 (0.71%) | 0.95 (0.94,0.96) | 0.2 (0.16,0.24) | 27.86 (22.81,33.43) | 0.04 (0.04,0.05) | 1.0 (1.00,1.00) | 0.94 (0.92,0.97) | 0.85 (0.84,0.85) | 6.13 (5.91,6.34) | 0.07 (0.04,0.10) |
| Mitral Regurgitation | 1368 (3.15%) | 0.92 (0.91,0.92) | 0.42 (0.39,0.45) | 13.46 (12.54,14.45) | 0.14 (0.13,0.15) | 0.99 (0.99,0.99) | 0.85 (0.83,0.87) | 0.83 (0.82,0.83) | 4.91 (4.76,5.06) | 0.18 (0.16,0.20) |
| RV Hypertension | 4300 (9.91%) | 0.88 (0.87,0.88) | 0.47 (0.45,0.48) | 4.7 (4.54,4.86) | 0.28 (0.27,0.29) | 0.98 (0.98,0.98) | 0.84 (0.83,0.85) | 0.77 (0.76,0.77) | 3.57 (3.49,3.65) | 0.21 (0.20,0.23) |
| Tricuspid Regurgitation | 2129 (4.91%) | 0.9 (0.89,0.90) | 0.39 (0.37,0.42) | 8.06 (7.62,8.53) | 0.17 (0.17,0.18) | 0.99 (0.99,0.99) | 0.84 (0.82,0.86) | 0.79 (0.79,0.80) | 4.05 (3.95,4.16) | 0.2 (0.18,0.22) |
| LV Systolic Dysfunction | 1831 (4.22%) | 0.94 (0.93,0.94) | 0.57 (0.55,0.60) | 13.57 (12.89,14.28) | 0.23 (0.22,0.24) | 0.99 (0.99,0.99) | 0.86 (0.85,0.88) | 0.88 (0.87,0.88) | 6.95 (6.72,7.17) | 0.16 (0.14,0.18) |
| Pulmonary Regurgitation | 1382 (3.18%) | 0.93 (0.92,0.93) | 0.49 (0.46,0.52) | 15.34 (14.35,16.42) | 0.15 (0.15,0.16) | 0.99 (0.99,0.99) | 0.84 (0.82,0.86) | 0.85 (0.84,0.85) | 5.48 (5.31,5.66) | 0.18 (0.16,0.21) |
| LV Hypertrophy | 608 (1.40%) | 0.88 (0.86,0.89) | 0.12 (0.10,0.14) | 8.69 (7.52,9.91) | 0.05 (0.05,0.06) | 1.0 (1.00,1.00) | 0.82 (0.79,0.86) | 0.79 (0.79,0.80) | 4.01 (3.84,4.19) | 0.22 (0.18,0.26) |
| RVOTO | 1242 (2.86%) | 0.88 (0.88,0.89) | 0.2 (0.18,0.22) | 6.92 (6.32,7.58) | 0.11 (0.10,0.11) | 0.99 (0.99,0.99) | 0.79 (0.76,0.81) | 0.81 (0.81,0.81) | 4.13 (3.99,4.28) | 0.26 (0.23,0.29) |

**Table S2: External Performance Metrics for Qualitative Outcomes**

| Outcome | N (%) | AUROC | AUPRC | Lift | PPV | NPV | Sensitivity | Specificity | LR+ | LR- |
|---|---|---|---|---|---|---|---|---|---|---|
| LVOTO | 89 (2.87%) | 0.79 (0.75,0.83) | 0.12 (0.07,0.17) | 4.12 (2.66,6.06) | 0.07 (0.05,0.08) | 0.99 (0.98,0.99) | 0.72 (0.62,0.81) | 0.71 (0.69,0.72) | 2.44 (2.10,2.79) | 0.4 (0.28,0.54) |
| Aortic Regurgitation | 135 (4.36%) | 0.8 (0.76,0.84) | 0.31 (0.23,0.40) | 7.2 (5.38,9.30) | 0.17 (0.14,0.21) | 0.98 (0.97,0.98) | 0.58 (0.49,0.66) | 0.88 (0.86,0.89) | 4.65 (3.84,5.49) | 0.48 (0.38,0.59) |
| Aortic Stenosis | 37 (1.20%) | 0.86 (0.80,0.92) | 0.13 (0.07,0.22) | 11.02 (5.96,18.55) | 0.04 (0.03,0.06) | 1.0 (0.99,1.00) | 0.76 (0.61,0.89) | 0.79 (0.77,0.80) | 3.55 (2.80,4.24) | 0.31 (0.13,0.50) |
| Mitral Regurgitation | 227 (7.33%) | 0.8 (0.77,0.83) | 0.34 (0.28,0.41) | 4.63 (3.81,5.54) | 0.27 (0.23,0.32) | 0.96 (0.95,0.97) | 0.5 (0.44,0.57) | 0.89 (0.88,0.90) | 4.75 (3.95,5.54) | 0.56 (0.48,0.63) |
| RV Hypertension | 256 (8.27%) | 0.74 (0.70,0.77) | 0.2 (0.16,0.24) | 2.4 (2.01,2.83) | 0.19 (0.17,0.22) | 0.95 (0.94,0.96) | 0.54 (0.48,0.60) | 0.8 (0.78,0.81) | 2.68 (2.31,3.06) | 0.58 (0.50,0.65) |
| Tricuspid Regurgitation | 220 (7.11%) | 0.73 (0.70,0.77) | 0.21 (0.16,0.26) | 2.96 (2.34,3.68) | 0.17 (0.14,0.21) | 0.95 (0.94,0.96) | 0.44 (0.37,0.50) | 0.84 (0.83,0.85) | 2.76 (2.30,3.26) | 0.67 (0.59,0.75) |
| LV Systolic Dysfunction | 58 (1.87%) | 0.85 (0.80,0.89) | 0.19 (0.10,0.29) | 10.33 (5.71,16.09) | 0.09 (0.06,0.13) | 0.99 (0.99,0.99) | 0.5 (0.38,0.63) | 0.91 (0.90,0.92) | 5.52 (4.09,7.18) | 0.55 (0.41,0.68) |
| Pulmonary Regurgitation | 105 (3.39%) | 0.73 (0.67,0.78) | 0.18 (0.11,0.26) | 5.38 (3.41,7.72) | 0.12 (0.08,0.16) | 0.97 (0.97,0.98) | 0.31 (0.23,0.40) | 0.92 (0.91,0.93) | 3.83 (2.71,5.06) | 0.75 (0.65,0.84) |
| LV Hypertrophy | 62 (2.00%) | 0.84 (0.79,0.89) | 0.13 (0.08,0.19) | 6.45 (4.28,9.79) | 0.07 (0.05,0.09) | 0.99 (0.99,1.00) | 0.69 (0.57,0.80) | 0.81 (0.79,0.82) | 3.6 (2.95,4.24) | 0.38 (0.24,0.53) |
| RVOTO | 172 (5.56%) | 0.75 (0.72,0.78) | 0.15 (0.11,0.19) | 2.67 (2.10,3.35) | 0.12 (0.10,0.15) | 0.96 (0.96,0.97) | 0.51 (0.43,0.58) | 0.79 (0.78,0.81) | 2.42 (2.04,2.80) | 0.62 (0.53,0.72) |

**Table S3: Aortic and Mitral Regurgitation Subgroup Analysis**

| | Aortic Regurgitation | | Mitral Regurgitation | |
|---|---|---|---|---|
| | Internal | External | Internal | External |
| Overall Cohort | 0.94 (0.93,0.95) | 0.8 (0.76,0.84) | 0.92 (0.91,0.92) | 0.8 (0.77,0.83) |
| Age Subgroups (years) | | | | |
| <1 | 0.87 (0.82,0.91) | 0.82 (0.72,0.90) | 0.91 (0.90,0.93) | 0.77 (0.72,0.83) |
| 1-3 | 0.9 (0.84,0.95) | — | 0.89 (0.87,0.91) | 0.79 (0.70,0.86) |
| 3-8 | 0.96 (0.93,0.98) | 0.79 (0.70,0.87) | 0.93 (0.91,0.94) | 0.8 (0.74,0.87) |
| 8-12 | 0.96 (0.93,0.98) | 0.76 (0.64,0.87) | 0.93 (0.91,0.95) | 0.76 (0.65,0.87) |
| 12-18 | 0.96 (0.95,0.97) | 0.77 (0.68,0.86) | 0.95 (0.94,0.96) | 0.87 (0.76,0.96) |
| ≥18 | 0.91 (0.89,0.93) | 0.81 (0.66,0.95) | 0.87 (0.85,0.90) | 0.79 (0.67,0.89) |
| CHD Subgroups | | | | |
| Composite Non-Critical CHD | 0.95 (0.92,0.97) | 0.82 (0.73,0.89) | 0.88 (0.86,0.90) | 0.76 (0.68,0.84) |
| Composite Critical CHD | 0.91 (0.86,0.95) | — | 0.82 (0.79,0.84) | 0.71 (0.61,0.81) |
| Any CHD | 0.94 (0.92,0.96) | 0.78 (0.70,0.86) | 0.88 (0.87,0.90) | 0.77 (0.70,0.83) |
| No CHD | 0.94 (0.89,0.97) | 0.77 (0.61,0.92) | 0.93 (0.91,0.95) | 0.89 (0.84,0.93) |
| Any Cardiomyopathy | 0.95 (0.91,0.98) | — | 0.91 (0.89,0.93) | 0.9 (0.76,1.00) |

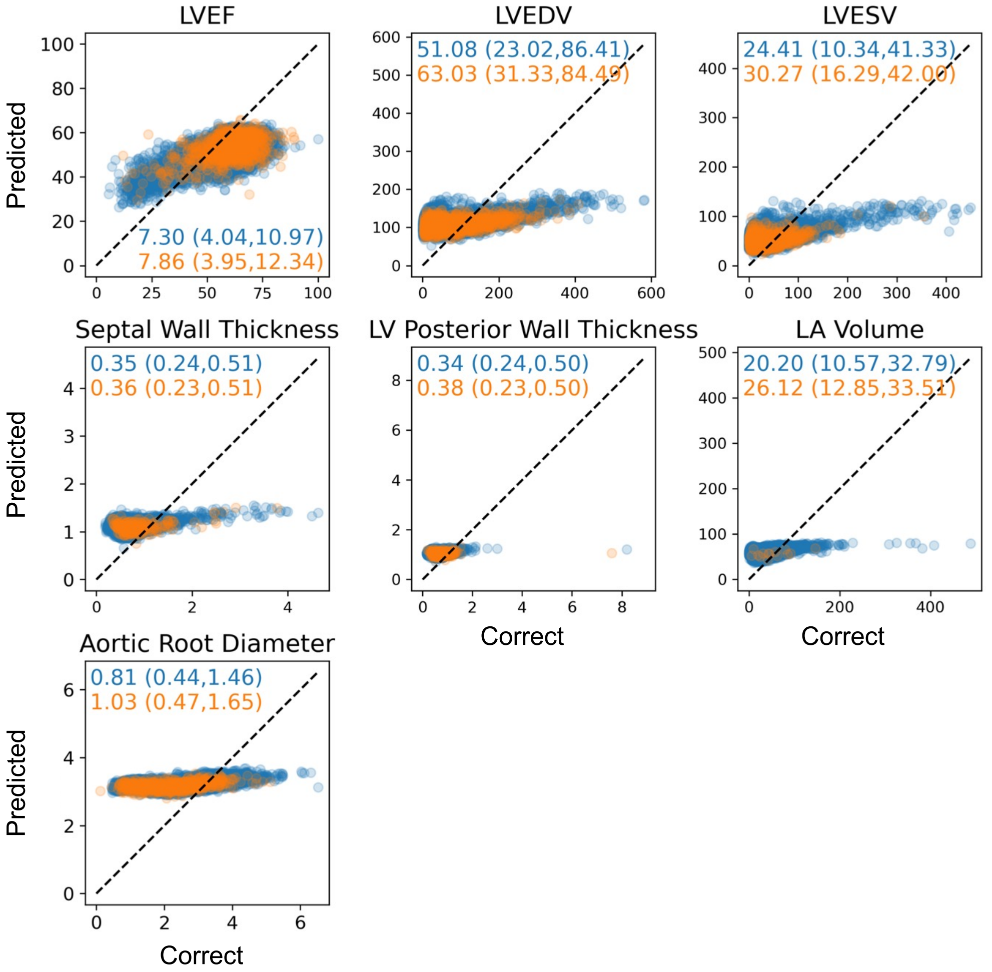

**Figure S1: Benchmark PanEcho Regression Task Performance.** Internal (blue) and external (orange) performance of the benchmark PanEcho to predict 7 measurements in the pediatric and CHD population. MAE values inset. Dotted line represents the identity line.

**Abbreviations:** left ventricular ejection fraction (LVEF); LV end-diastolic volume (LVEDV); LV end-systolic volume (LVESV); left atrium (LA).
